# Aspergillosis and Mucormycosis in COVID-19 Patients; a Systematic Review and Meta-analysis

**DOI:** 10.1101/2021.08.01.21261458

**Authors:** Saira Afzal, Mehreen Nasir

## Abstract

Fungal infections have increased in number since the onset of this lethal pandemic. The aim of this study is to assess risk factors and case fatality in COVID-19 cases with aspergillosis or mucormycosis. Systematic review and meta-analysis was done according to PRISMA guidelines. Data bases used were Google scholar, Pakmedinet, PUBMED and MEDLINE. 21 case reports and case series of mucormycosis in COVID-19 patients were identified and mean age was 56.3 years (36 males and 12 females). The most common comorbidity was diabetes and site was Rhino orbital mucormycosis. Case fatality of 48 combined cases was calculated to be 52%. 19 articles of aspergillosis were included. Diabetes was the most common comorbidity in cases. The number of male cases were more than females. Incidence of aspergillosis in critically sick COVID-19 patients was calculated to be 9.3%. Case fatality was calculated to be 51.2%. Screening can be a beneficial tool for decreasing the morbidity and mortality.

## INTRODUCTION

COVID-19 pandemic started in China and subsequently spread throughout the world at an alarming rate. This disease can involve multiple systems of the body. Recently, researchers and medical care providers have noticed that fungal infections such as aspergillosis and mucormycosis are on an increasing trend among patients already infected with this lethal covid virus. The spores of these fungi are spread everywhere in environment. Normal healthy people continue to breathe in the air without being affected. Getting infected with these fungi is a rare occurrence but patients who are already in an immunocompromised state and suffering from lung diseases due to Covid-19 are more prone to acquiring these devastating pathogens.

Mucormycosis is also known as black fungus. It can affect nasal cavity, sinuses, lungs, gastrointestinal tract and skin. When pathogen enters the blood stream it can cause disseminated mucormycosis. Rhino cerebral is the most common site involved by mucormycosis. This disease does not spread by person to person contact. Treatment is done with anti-fungal drugs and most cases do require surgical recession of site involved.^1^ Aspergillosis commonly known as mold also affects those who have weakened immune system or any lung disease for example asthmatics or chronic obstructive lung disease. The mortality with this disease is high.^2^ Aspergillosis and mucormycosis have high prevalence covid-19 positive population. A study done on 184,500,000 people has revealed that 1.78% have shown to have serious fungal infections.^3^ Pakistan has high burden of non-communicable diseases. Surveillance of non-communicable diseases is limited. People are underdiagnosed which causes complications as these diseases are not managed. Fungal infections are opportunistic infections which have high incidence in people with preexisting diseases. Secondly, Pakistan has a high prevalence of tuberculosis which is a risk factor as well.^4-^ COVID-19 is a respiratory disease which is a risk factor for opportunistic infection. Influenza, a respiratory illness has also shown to increase the incidence of aspergillosis.^5^

The rationale of this study is to identify the risk factors and case fatality of aspergillosis or mucormycosis in COVID-19 cases. This study will also identify the incidence of aspergillosis in critically sick persons that will include both ICU patients and mechanically ventilated cases. Knowledge generated from this systematic review will highlight the importance of preventing and early screening in critically sick persons of COVID-19 as this disease has high morbidity and mortality.

## METHODOLOGY

Systematic review was done according to PRISMA guidelines. My inclusion criteria was original articles and abstracts on aspergillosis or mucormycosis in COVID-19 patients from 1^st^ January 2020 to 15^th^ June, 2021.

Articles focusing on treatment, management, surgical options, autopsy, postmortem, histopathology findings, molecular detection, genetic studies and laboratory reports were excluded. Articles with language other than English were also excluded.

Data bases used were Google scholar, Pakmedinet, PUBMED and MEDLINE. PRISMA flowchart is shown in the figure 1. For finding articles Boolean operators of ‘COVID-19 AND (ASPERGILLOSIS OR MUCORMYCOSIS)’ were used. Date of my search was 15^th^ June, 2021.

**Figure 1:**
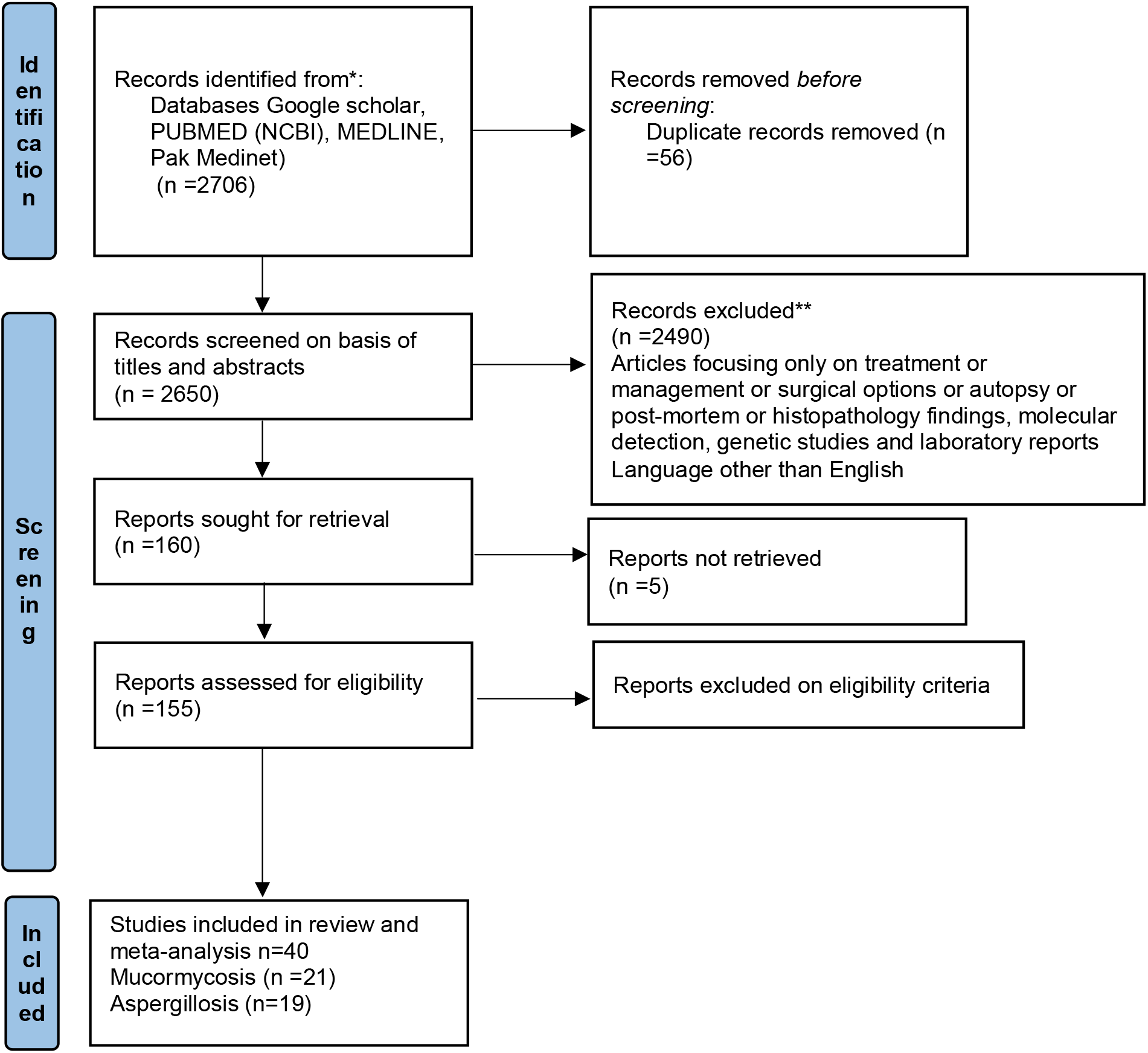
Identification of studies via databases and registers.

**Figure 2:**
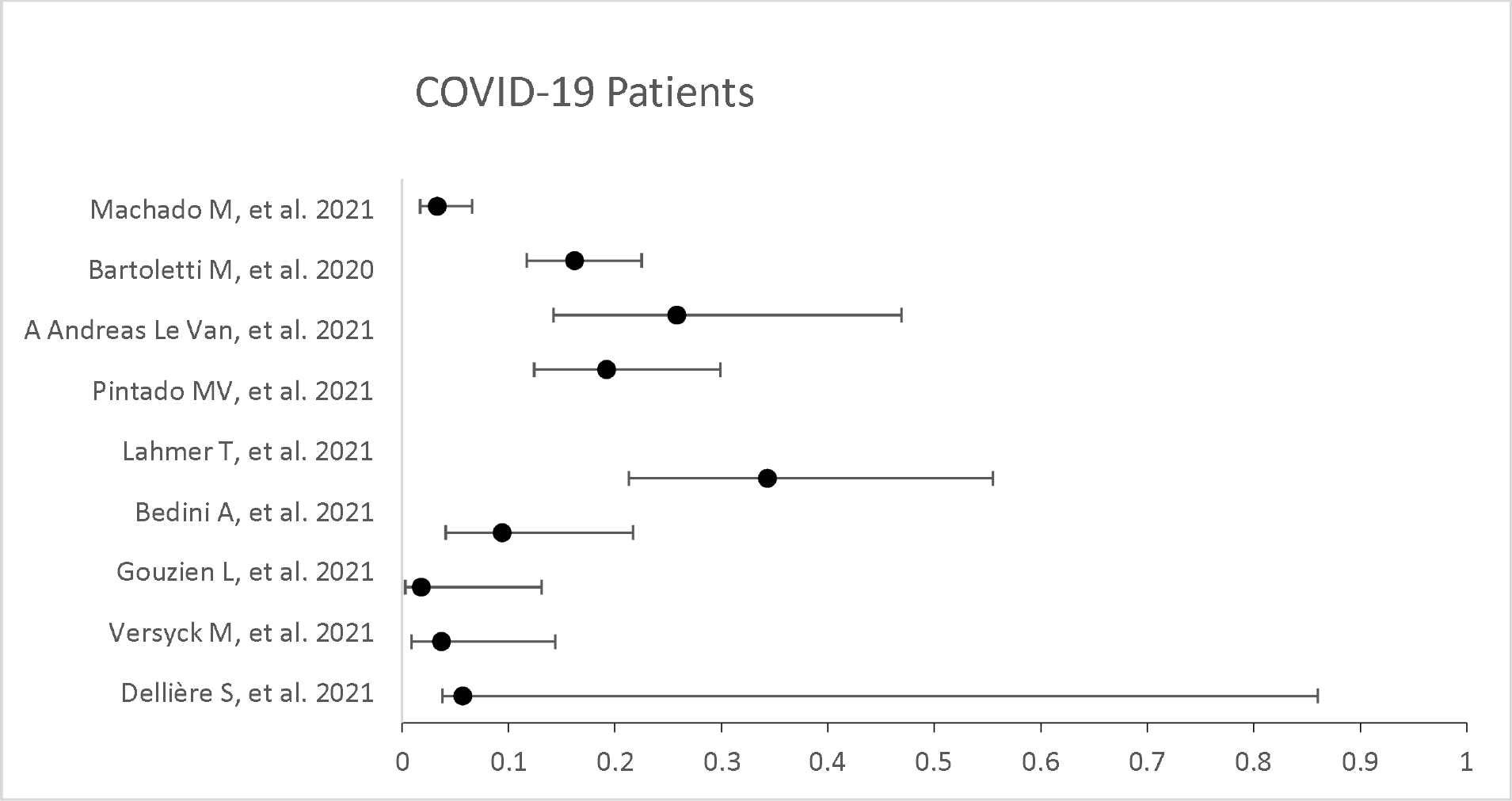
Incidence Rate Ratio of Aspergillosis in critically sick.

2 researchers independently screened the articles of basis of title and abstract. Any discrepancy was solved by discussion. Data of outcome variables was entered in Microsoft Excel. For meta-analysis of mucormycosis in COVID-19 cases 21 case reports and case series were included. Age, gender, site of mucormycosis, preexisting co-morbidities and outcome was noted. For aspergillosis in COVID-19; 19 articles were selected. Case fatality was calculated. Incidence of aspergillosis in COVID-19 patients was calculated. Case reports were excluded for doing meta-analysis of aspergillosis.

## RESULTS

21 case reports and case series of mucormycosis in COVID-19 patients were identified as shown in table I. There were total 48 patients in analysis. Mean age was found to be 56.312 years with standard deviation of 15.55 years. There were 36 males and 12 females. The most common comorbidity was diabetes (35 out of 48 cases). Other risk factors included chronic kidney disease, ischemic heart disease, transplant, decompensated liver disease and hypertension. The most common site was Rhino orbital mucormycosis. However cutaneous, gastrointestinal and pulmonary mucormycosis were also reported. 25 cases died and 23 were discharged. Chi-square test did not show significant difference in outcome between males and females (p-value 0.133). Combined case fatality of 48 cases was calculated to be 52%.

**Table I:**
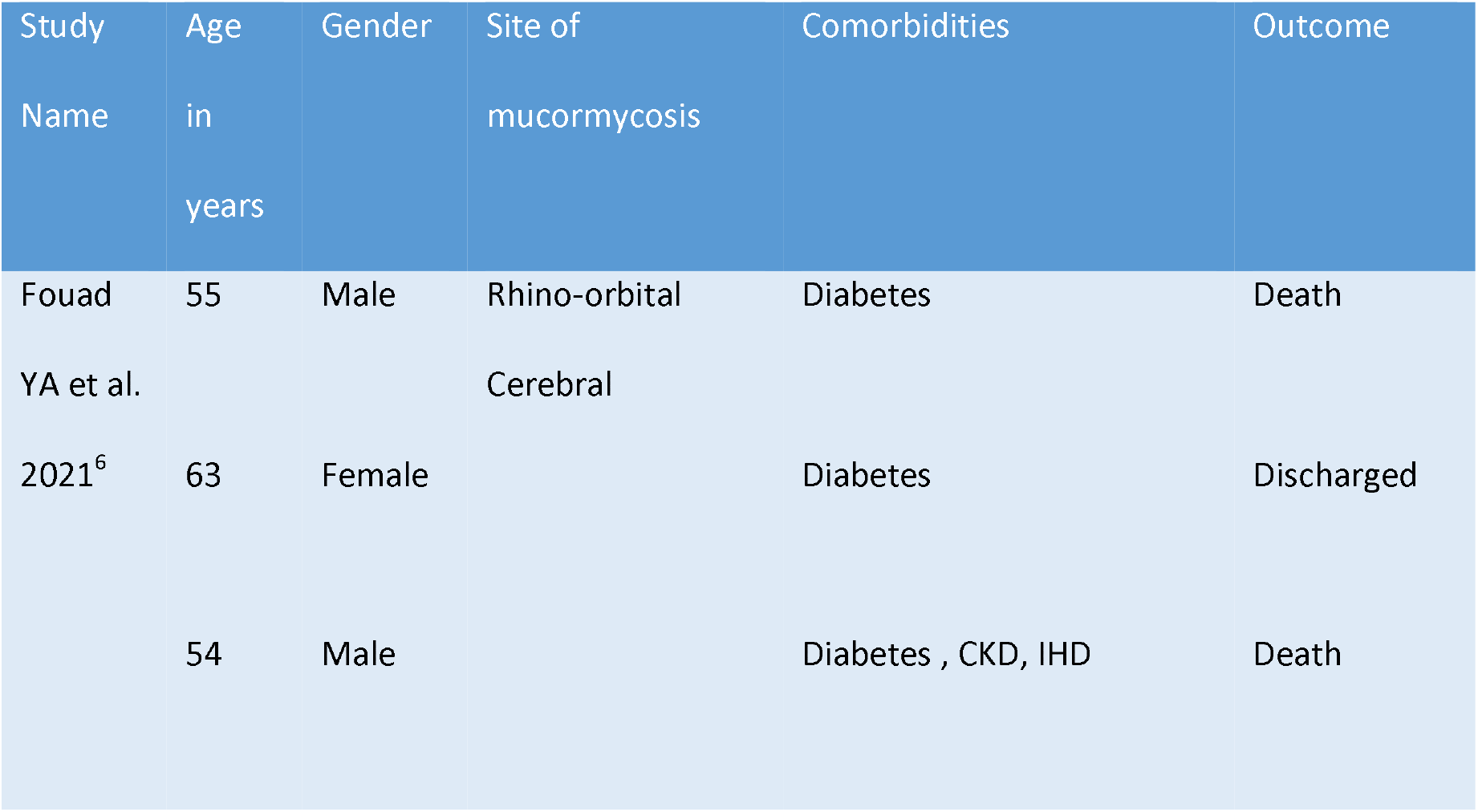
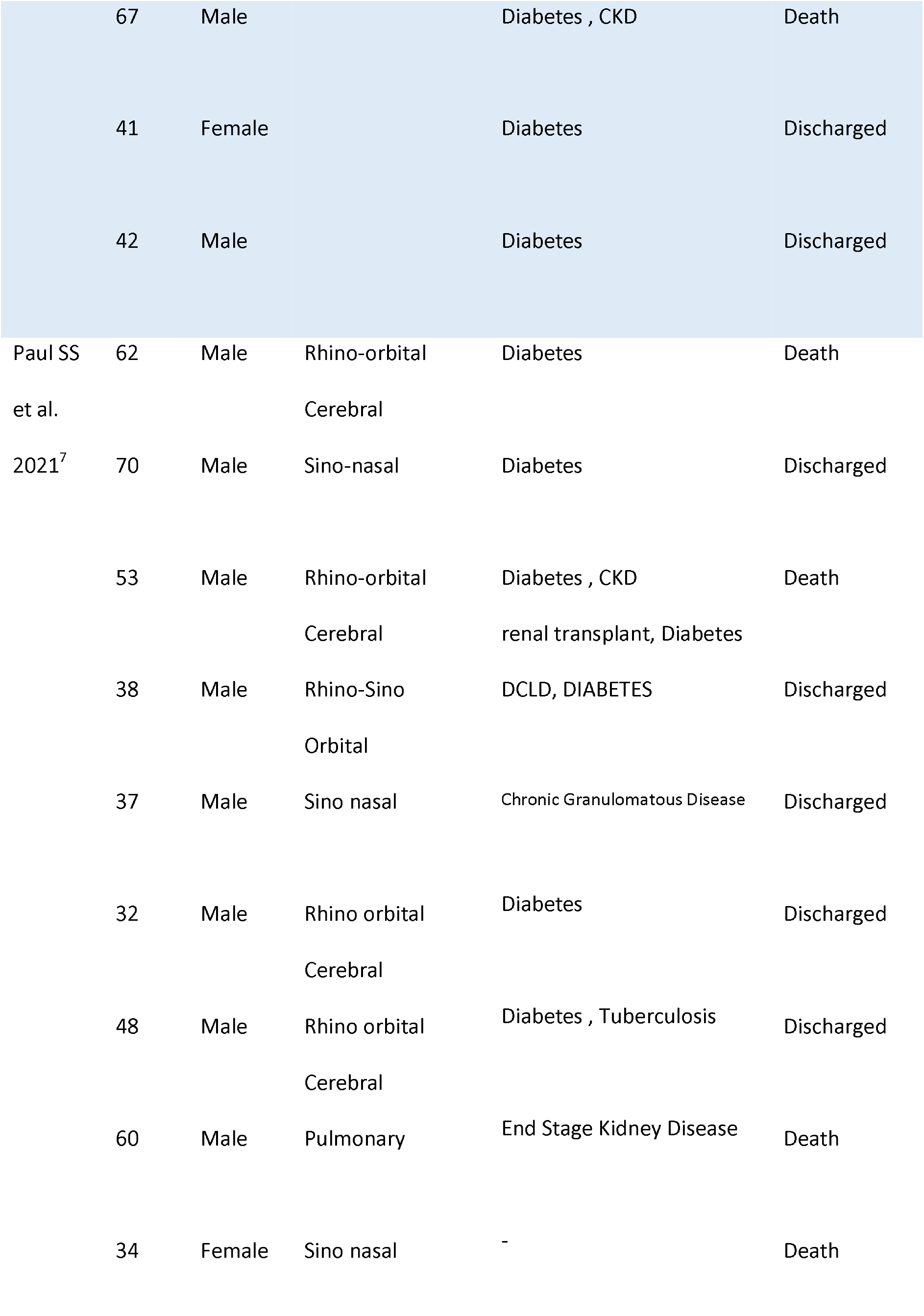
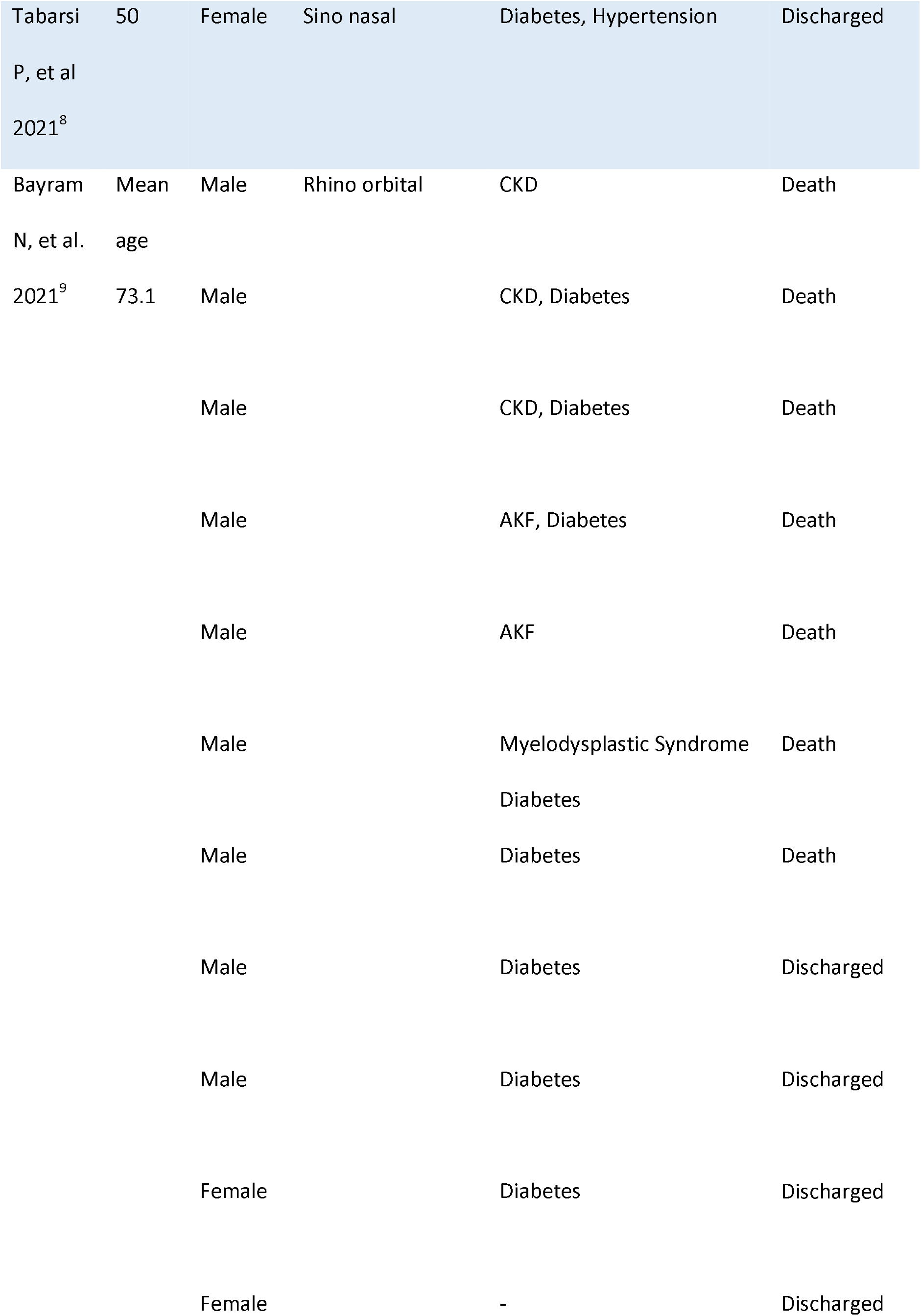
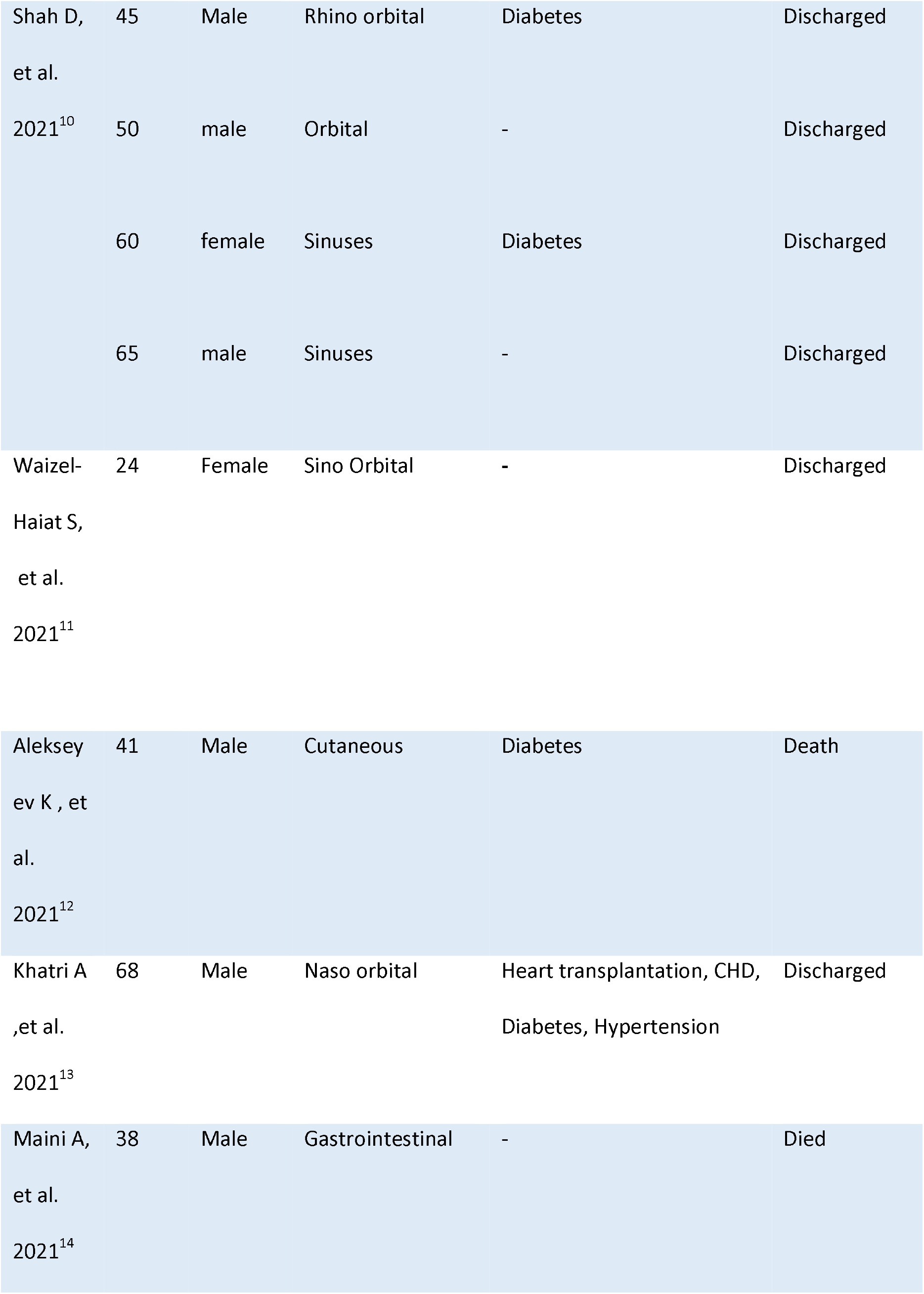
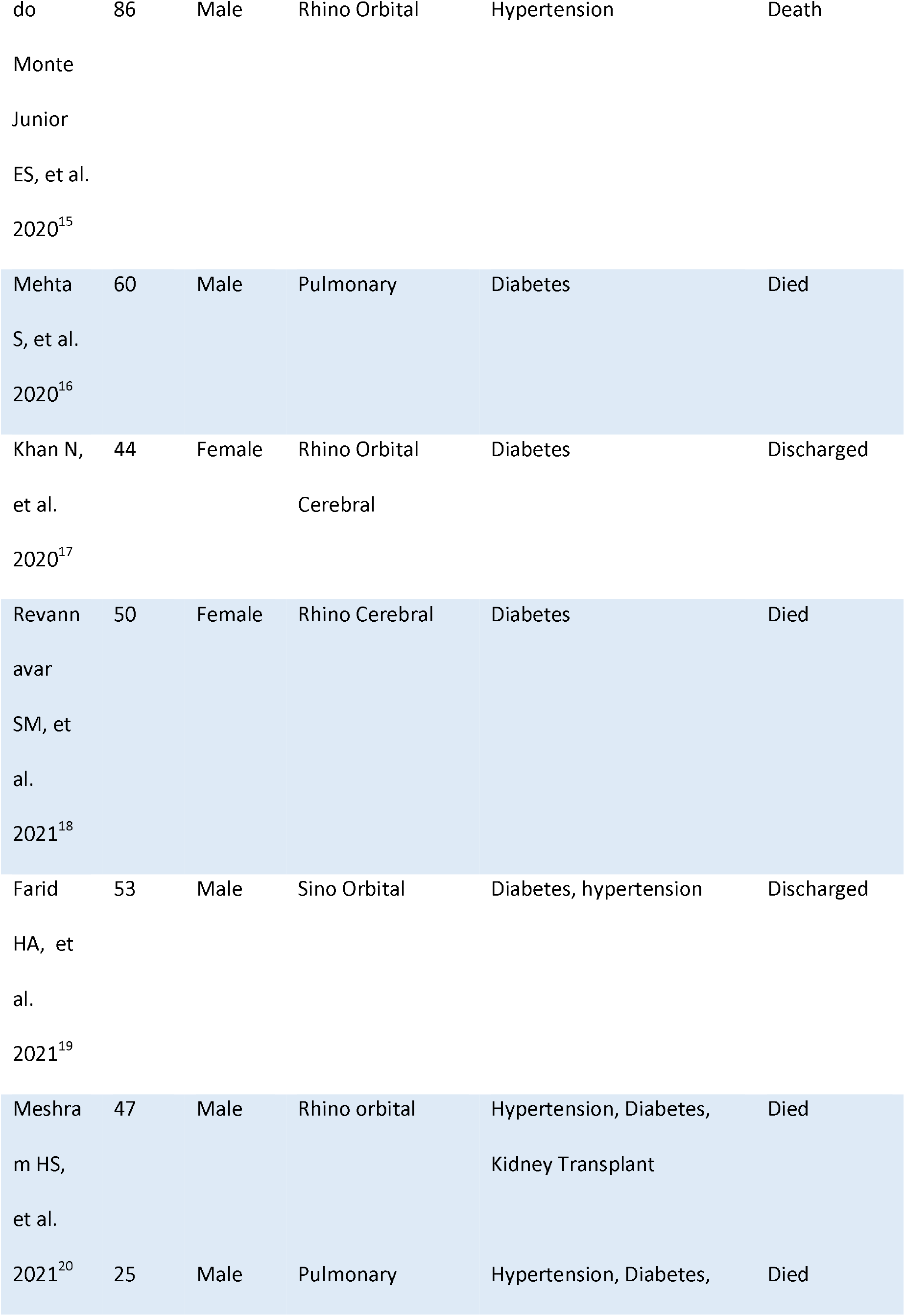
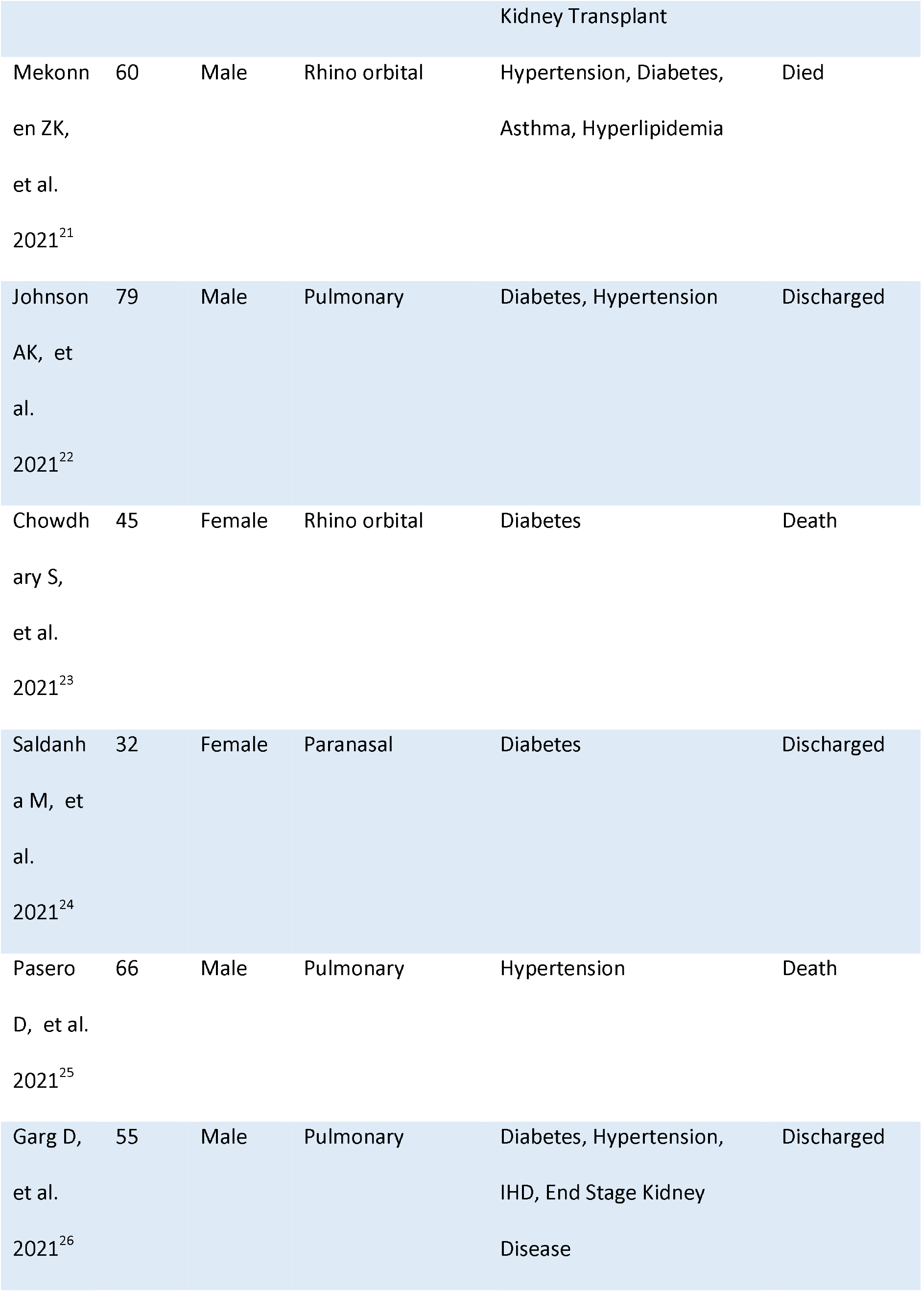
Case Reports and Case Series of Mucormycosis.

19 articles meeting the inclusion criteria were included. Diabetes was the most common comorbidity in cases. Other risk factors include hypertension, steroid use, hypertension, heart disease, chronic obstructive pulmonary disease, malignancies and chronic kidney disease. The number of male cases were more than females.^36,40^ Combined incidence was 9.3%. Case fatality was calculated to be 51.2% as shown in Table II. Incidence of aspergillosis in critically sick COVID-19 patients was calculated by including 9 studies as shown in Table III. Forest plot shows the incidence of aspergillosis in critically sick COVID-19 patients using 95% confidence interval.

**Table II:** Number of cases who died due to aspergillosis in COVID-19.

| Study | Total number of cases N | Number of cases who died n (%) |
| --- | --- | --- |
| Nasir N, et al. 2021<sup>36</sup> | 9 | 4 (44.4%) |
| Lai CC, et al. 2021<sup>37</sup> | 34 | 22 (64.7%) |
| Kariyawasam RM, et al. 2021<sup>38</sup> | 182 | 91 (50%) |
| Chong WH, et al. 2021<sup>39</sup> | 192 | 94 (48.9%) |
| Pintado MV, et al. 2021<sup>40</sup> | 16 | 5 (31.2%) |
| Apostolopoulou A, et al. 2021<sup>41</sup> | 85 | 46 (54.1%) |
| Salmanton-Garcia J, et al. 2021<sup>42</sup> | 186 | 97 (19.9%) |
| Abdalla S, et al. 2021<sup>43</sup> | 2 | 2 (100%) |
| Alobaid K, et al. 2021<sup>44</sup> | 2 | 2 (100%) |
| Total | 708 | 363 (51.2%) |

**Table III:**
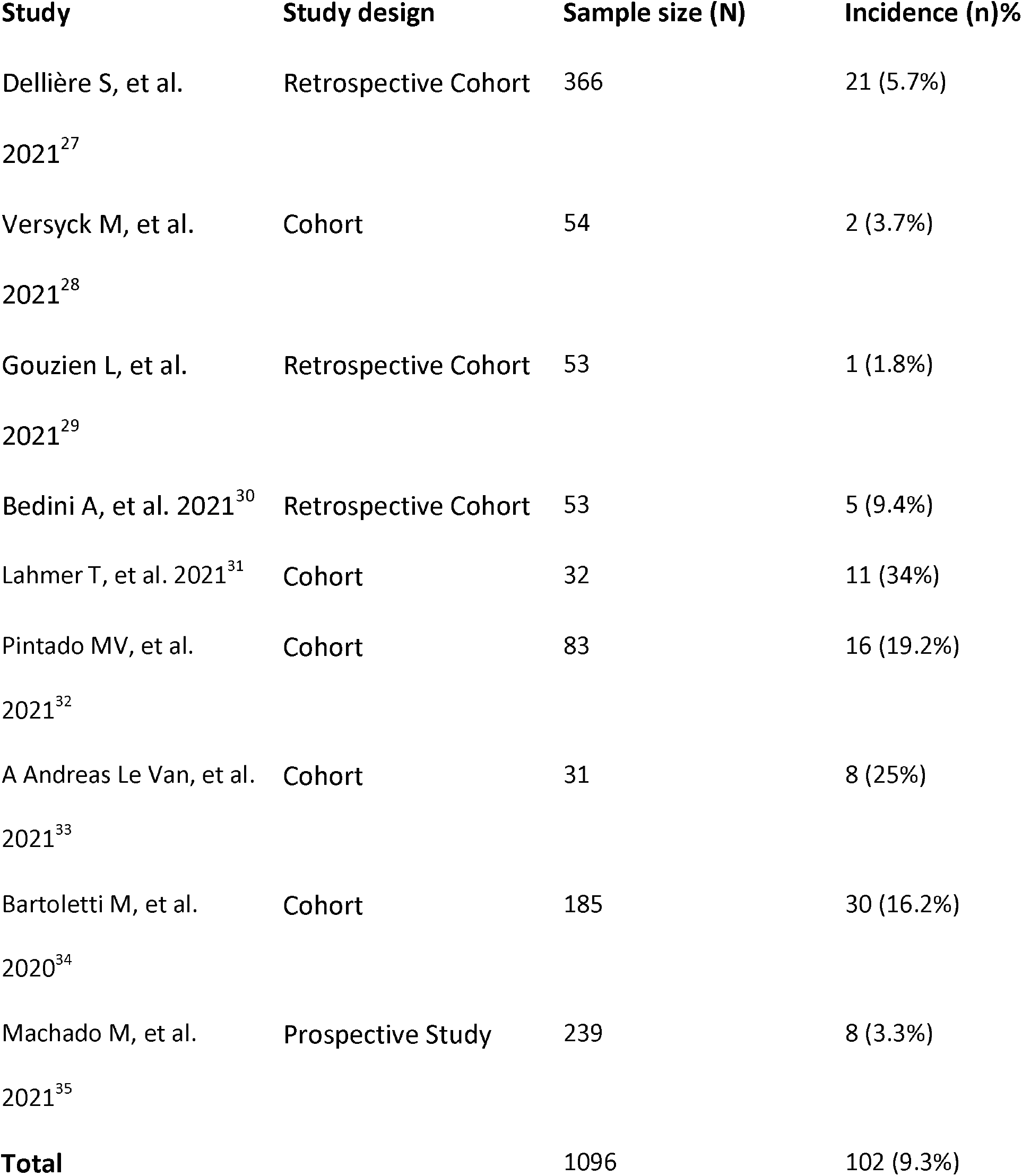
Incidence of aspergillosis in critically sick patients of COVID-19.

| Study | Study design | Sample size (N) | Incidence (n)% |
| --- | --- | --- | --- |
| Dellière S, et al.<br>2021<sup>27</sup> | Retrospective Cohort | 366 | 21 (5.7%) |
| Versyck M, et al.<br>2021<sup>28</sup> | Cohort | 54 | 2 (3.7%) |
| Gouzien L, et al.<br>2021<sup>29</sup> | Retrospective Cohort | 53 | 1 (1.8%) |
| Bedini A, et al. 2021<sup>30</sup> | Retrospective Cohort | 53 | 5 (9.4%) |
| Lahmer T, et al. 2021<sup>31</sup> | Cohort | 32 | 11 (34%) |
| Pintado MV, et al.<br>2021<sup>32</sup> | Cohort | 83 | 16 (19.2%) |
| A Andreas Le Van, et al.<br>2021<sup>33</sup> | Cohort | 31 | 8 (25%) |
| Bartoletti M, et al.<br>2020<sup>34</sup> | Cohort | 185 | 30 (16.2%) |
| Machado M, et al.<br>2021<sup>35</sup> | Prospective Study | 239 | 8 (3.3%) |
| **Total** |  | 1096 | 102 (9.3%) |

## Discussion

The aim of this study was to assess the risk factors and case fatality of aspergillosis and mucormycosis in COVID-19 patients. Incidence of aspergillosis in critically sick patients that included both intensive care unit patients and mechanically ventilated patients of COVID-19 has also been calculated. This study highlighted the importance of prevention and early screening of fungal infections in severe cases of COVID-19 as this lethal disease has high morbidity and mortality. Repeated surgical interventions are required to remove infected tissue which causes disfigurement and mental trauma.

Mean age of cases with mucormycosis was 56 years. This result is similar to other studies.^45^ The number of males are more than females. This finding is consistent with previous study.^46^ A retrospective study in Mexico has shown more male patients of mucormycosis. There are more males in this study as severe COVID-19 and mucormycosis both are generally more common in males.^47^

Diabetes was the most common risk factor found in other systematic reviews of mucormycosis.^48-95^ Diabetes is an immunocompromised state and its prevalence is high across the globe. Rhino orbital mucormycosis was the most common site in cases.^50^ The combined case fatality in this study was found to be 52%. The mortality of pulmonary mucormycosis is slightly higher. A study done has demonstrated mortality of 57%. Another systematic review on mucormycosis has shown 67% survival with antifungal medications and surgery.^51^ This study has reported higher mortality (52%). This may be due to the fact that cases of mucormycosis has COVID-19 infection as well which has resulted in higher case fatality.

Aspergillosis has higher incidence in critically sick COVID-19 patients. That demands for early screening. The combined incidence was found to be 9.3% with range of 1.88% to 25%. In another systematic review the incidence was found to be 13.5%.^51^ This may be due to the fact that in this study both mechanically ventilated and not mechanically ventilated Intensive Care Unit (ICU) patients were included. The average case fatality was found to be 51.2%. Another systematic review has calculated the case fatality of aspergillosis to be 58%.^52^ A systematic review of pulmonary aspergillosis in COVID-19 cases; case fatality was found to be 48.4%.^51^ The risk factors of aspergillosis were diabetes, hypertension, steroid use, hypertension, heart disease, chronic obstructive pulmonary disease, malignancies and chronic kidney disease.

The strengths of this study are that it is a systematic review and meta-analysis. Thorough data search has been done. It has covered 2 lethal fungal infections which have shown increased prevalence since covid pandemic. However more prospective studies should be done to understand more about pathogenesis of this disease. The most common risk factor was diabetes. Health education and lifestyle modifications should be promoted to halt the rise in diabetes. Early screening and good glycemic control can also be helpful for those who have already developed diabetes to prevent complications.

## Conclusion

Case fatality from aspergillosis and mucormycosis in Covid-19 cases is quite high. Incidence of aspergillosis in critically sick COVID cases is around 9.2%. Screening can be a beneficial tool for decreasing the morbidity and mortality.

## Data Availability

Data was collected from articles case reports series and prospective studies.

## Conflict Of Interest

None

## DISCLOSURE

None

## FUNDING

None

## Notes

### Competing Interest Statement

The authors have declared no competing interest.

### Funding Statement

No funding was done

### Author Declarations

This is a systematic Review

## References

1. Mucormycosis. Centers for Disease Control and Prevention. https://www.cdc.gov/fungal/diseases/mucormycosis/index.html. (Accessed 22/06/2021).

2. Aspergillosis. Centers for Disease Control and Prevention. https://www.cdc.gov/fungal/diseases/aspergillosis/index.html. (Accessed 22/06/2021).

3. Jabeen K, Farooqi J, Mirza S, Denning D, Zafar A. Serious fungal infections in Pakistan. Eur. J. Clin. Microbiol. Infect. Dis. 2017;36(6):949–56.

4. National Institute of Health Islamabad. National TB Control Program - National Institute of Health Islamabad. https://www.nih.org.pk/national-tb-control-program. (Accessed 22/06/2021).

5. Waldeck F, Boroli F, Suh N, Wendel Garcia PD, Flury D, Notter J, et al. Influenza-associated aspergillosis in critically-ill patients—a retrospective bicentric cohort study. Eur. J. Clin. Microbiol. Infect. Dis. 2020;39:1915–23.

6. Fouad YA, Abdelaziz TT, Askoura A, Saleh MI, Mahmoud MS, Ashour MM. Spike in rhino-orbital-cerebral mucormycosis cases presenting to a tertiary care center during the COVID-19 pandemic. Front. Med. 2021;8.

7. Clinical Characteristics And Outcomes of 16 Cases With COVID19 and Mucormycosis: Experience From A Tertiary Care Center In India and Review of Literature. Available from: https://assets.researchsquare.com/files/rs-533347/v1/78a6c40d-83f5-4c53-a31b-ef3c8a06a4bb.pdf [cited 15 July 2021].

8. COVID-19 associated rhinosinusitis mucormycosis due to Rhizopus arrhizus: A rare but potentially fatal infection occurring after treatment with corticosteroids. Available from: https://assets.researchsquare.com/files/rs-398594/v1/5e4c735f-f595-4a7e-a8f9-b76a3210c58c.pdf [cited 15 July 2021].

9. Bayram N, Ozsaygılı C, Sav H, Tekin Y, Gundogan M, Pangal E, et al. Susceptibility of severe COVID-19 patients to rhino-orbital mucormycosis fungal infection in different clinical manifestations. Jpn. J. Ophthalmol 2021;31:1–1.

10. Shah D, Talwar D, Kumar S, Acharya S, Dubey A. Mucormycosis as a complication of LONG COVID: A case series. J. Med. Sci. 2021;25(112):1331–7.

11. Waizel-Haiat S, Guerrero-Paz JA, Sanchez-Hurtado L, Calleja-Alarcon S, Romero-Gutierrez L. A case of fatal rhino-orbital mucormycosis associated with new onset diabetic ketoacidosis and COVID-19. Cureus 2021;13(2).

12. Alekseyev K, Didenko L, Chaudhry B. Rhinocerebral mucormycosis and COVID-19 pneumonia. J. Med. Case Rep. 2021;12(3):85.

13. Khatri A, Chang KM, Berlinrut I, Wallach F. Mucormycosis after Coronavirus disease 2019 infection in a heart transplant recipient–case report and review of literature. Med. Mycol. 2021;31(2):101125.

14. Maini A, Tomar G, Khanna D, Kini Y, Mehta H, Bhagyasree V. Sino-orbital mucormycosis in a COVID-19 patient: A case report. Int. J. Surg. Case Rep. 2021;82:105957.

15. do Monte Junior ES, Dos Santos ME, Ribeiro IB, de Oliveira Luz G, Baba ER, Hirsch BS, et al. Rare and Fatal Gastrointestinal Mucormycosis (Zygomycosis) in a COVID-19 Patient: A Case Report. Clin. Endosc. 2020;53(6):746.

16. Mehta S, Pandey A. Rhino-orbital mucormycosis associated with COVID-19. Cureus 2020;12(9).

17. Khan N, Gutierrez CG, Martinez DV, Proud KC. A case report of COVID-19 associated pulmonary mucormycosis. Arch. Clin. Cases. 2021;7(3):2020–7.

18. Revannavar SM, Supriya PS, Samaga L, Vineeth VK. COVID-19 triggering mucormycosis in a susceptible patient: a new phenomenon in the developing world?. BMJ Case Rep. 2021;14(4).

19. Farid HA, Hashim AR, Hasrat NH. Rhinocerebral Mucormycosis as a COVID-19-Related Complication: A Case Report from Basra City, Southern Iraq. Int. J. Glob. Sci. Res. 2021;6(5):1369–74.

20. Meshram HS, Kute VB, Chauhan S, Desai S. Mucormycosis in post-COVID-19 renal transplant patients: A lethal complication in follow-up. Transpl Infect Dis. 2021;3.

21. Mekonnen ZK, Ashraf DC, Jankowski T, Grob SR, Vagefi MR, Kersten RC, et al. Acute invasive rhino-orbital mucormycosis in a patient with COVID-19-associated acute respiratory distress syndrome. Ophthalmic Plast Reconstr Surg. 2021;37(2).

22. Johnson AK, Ghazarian Z, Cendrowski KD, Persichino JG. Pulmonary aspergillosis and mucormycosis in a patient with COVID-19. Med. Mycol. Case Rep. 2021;32:64–7.

23. Chowdhary S. The Face of the COVID-19 Pandemic: Invasive Rhino-Orbital Mucormycosis. Clin Surg J. 2021;4(S7):14–7.

24. Saldanha M, Reddy R, Vincent MJ. Paranasal mucormycosis in COVID-19 patient. Indian J. Otolaryngol. Head Neck Surg 2021;22:1–4.

25. Pasero D, Sanna S, Liperi C, Piredda D, Branca GP, Casadio L, et al. A challenging complication following SARS-CoV-2 infection: a case of pulmonary mucormycosis. Infection 2020 :1–6.

26. Garg D, Muthu V, Sehgal IS, Ramachandran R, Kaur H, Bhalla A, et al. Coronavirus disease (Covid-19) associated mucormycosis (CAM): case report and systematic review of literature. Mycopathologia 2021;5:1–0.

27. Dellière S, Dudoignon E, Fodil S, Voicu S, Collet M, Oillic PA, et al. Risk factors associated with COVID-19-associated pulmonary aspergillosis in ICU patients: a French multicentric retrospective cohort. Clin. Microbiol. Infect. 2021;27(5):790.

28. Versyck M, Zarrougui W, Lambiotte F, Elbeki N, Saint-Leger P. Invasive pulmonary aspergillosis in COVID-19 critically ill patients: Results of a French monocentric cohort. Med. Mycol. J. 2021;31(2):101122.

29. Gouzien L, Cocherie T, Eloy O, Legriel S, Bedos JP, Simon C, et al. Invasive Aspergillosis associated with Covid-19: A word of caution. Infectious Diseases Now 2021;51(4):383–6.

30. Bacterial and Fungal Co-Infections in Patients with COVID-19 Related Pneumonia: A Retrospective Cohort Study. Available from: https://www.researchsquare.com/article/rs-576997/v1 [cited 15 July 2021].

31. Lahmer T, Kriescher S, Herner A, Rothe K, Spinner CD, Schneider J, et al. Invasive pulmonary aspergillosis in critically ill patients with severe COVID-19 pneumonia: Results from the prospective AspCOVID-19 study. Plos one. 2021;16(3).

32. Pintado MV, Camiro-Zúñiga A, Soto MA, Cuenca D, Mercado M, Crabtree-Ramirez B. COVID-19-associated invasive pulmonary aspergillosis in a tertiary care center in Mexico City. Med Mycol. 2021.

33. van Arkel, A., Rijpstra, T. A., Belderbos, H., van Wijngaarden, P., Verweij, P. E., Bentvelsen R. G. COVID-19-associated Pulmonary Aspergillosis. Am J Respir Crit Care Med. 2020;202(1):132–135.

34. Bartoletti M, Pascale R, Cricca M, Rinaldi M, Maccaro A, Bussini L, et al. Epidemiology of invasive pulmonary aspergillosis among COVID-19 intubated patients: a prospective study. Clin Infect Dis. 2020.

35. Machado M, Valerio M, Àlvarez-Uría A, Olmedo M, Veintimilla C, Padilla B, et al. Invasive pulmonary aspergillosis in the COVID-19 era: An expected new entity. Mycoses 2021;64(2):132–43.

36. Nasir N, Farooqi J, Mahmood SF, Jabeen K. COVID-19-associated pulmonary aspergillosis (CAPA) in patients admitted with severe COVID-19 pneumonia: an observational study from Pakistan. Mycoses 2020;63(8):766–70.

37. Lai CC, Yu WL. COVID-19 associated with pulmonary aspergillosis: A literature review. J Microbiol Immunol Infect. 2021;54(1):46–53.

38. Kariyawasam RM, Dingle TC, Kula BE, Sligl WI, Schwartz IS. COVID-19 Associated Pulmonary Aspergillosis: Systematic Review and Patient-Level Meta-analysis. medRxiv. 2021.

39. Chong WH, Neu KP. The incidence, diagnosis, and outcomes of COVID-19-associated pulmonary aspergillosis (CAPA): a systematic review. J Hosp Infect. 2021;113: 115–129.

40. Pintado MV, Camiro-Zúñiga A, Soto MA, Cuenca D, Mercado M, Crabtree-Ramirez B. COVID-19-associated invasive pulmonary aspergillosis in a tertiary care center in Mexico City. Med mycol. 2021.

41. Apostolopoulou A, Esquer Garrigos Z, Vijayvargiya P, Lerner AH, Farmakiotis D. Invasive pulmonary aspergillosis in patients with SARS-CoV-2 infection: a systematic review of the literature. Diagnostics 2020;10(10):807.

42. Salmanton-Garcia J, Sprute R, Stemler J, Bartoletti M, Dupont D, Valerio M, et al. COVID-19–associated pulmonary aspergillosis, March–August 2020. Emerg. Infect. Dis. 2021;27(4):1077.

43. Abdalla S, Almaslamani MA, Hashim SM, Ibrahim AS, Omrani AS. Fatal coronavirus disease 2019-associated pulmonary aspergillosis; a report of two cases and review of the literature. IDCases. 2020;22.

44. Alobaid K, Yousuf B, Al-Qattan E, Muqeem Z, Al-Subaie N. Pulmonary aspergillosis in two COVID-19 patients from Kuwait. Access Microbiol 2021;3(3):000201.

45. Ravani SA, Agrawal GA, Leuva PA, Modi PH, Amin KD. Rise of the phoenix: Mucormycosis in COVID-19 times. Indian J. Ophthalmol. 2021;69(6):1563–8.

46. Pakdel F, Ahmadikia K, Salehi M, Tabari A, Jafari R, Mehrparvar G, et al. Mucormycosis in patients with COVID-19: A cross-sectional descriptive multicenter study from Iran. Mycoses 2021.

47. Bonifaz A, Tirado-Sánchez A, Hernández-Medel ML, Araiza J, Kassack JJ, del Angel-Arenas T, Moisés-Hernández JF, et al. Mucormycosis at a tertiary-care center in Mexico. A 35-year retrospective study of 214 cases. Mycoses 2021;64(4):372–80.

48. Jeong W, Keighley C, Wolfe R, Lee WL, Slavin MA, Kong DC, et al. The epidemiology and clinical manifestations of mucormycosis: a systematic review and meta-analysis of case reports. Clin. Microbiol. Infect. 2019;25(1):26–34.

49. Vaezi A, Moazeni M, Rahimi MT, de Hoog S, Badali H. Mucormycosis in Iran: a systematic review. Mycoses 2016;59(7):402–15.

50. Muthu V, Agarwal R, Dhooria S, Sehgal IS, Prasad KT, Aggarwal AN, et al. Has the mortality from pulmonary mucormycosis changed over time? a systematic review and meta-analysis. Clin. Microbiol. Infect. 2021;27(4):538–549.

51. Chong WH, Neu KP. The incidence, diagnosis, and outcomes of COVID-19-associated pulmonary aspergillosis (CAPA): a systematic review. J Hosp Infect. 2021;113:115–129.

52. Lin SJ, Schranz J, Teutsch SM. Aspergillosis case-fatality rate: systematic review of the literature. Clin. Infect. Dis. 2001;32(3):358–66.

